# How deadly is COVID-19? A rigorous analysis of excess mortality and age-dependent fatality rates in Italy

**DOI:** 10.1101/2020.04.15.20067074

**Authors:** Chirag Modi, Vanessa Böhm, Simone Ferraro, George Stein, Uroš Seljak

## Abstract

We perform a counterfactual time series analysis on 2020 mortality data from towns in Italy using data from the previous five years as control. We find an excess mortality that is correlated in time with the official COVID-19 death rate, but exceeds it by a factor of at least 1.5. Our analysis suggests that there is a large population of predominantly older people that are missing from the official fatality statistics. We estimate that the number of cOvID-19 deaths in Italy is 49,000-53,000 as of May 9 2020, as compared to the official number of 33,000. The Population Fatality Rate (PFR) has reached 0.26% in the most affected region of Lombardia and 0.58% in the most affected province of Bergamo. These PFRs constitutes a lower bound to the Infection Fatality Rate (IFR). We combine the PFRs with the Test Positivity Ratio to derive the lower bound of 0.61% on the IFR for Lombardia. We further estimate the IFR as a function of age and find a steeper age dependence than previous studies: we find 17% of COVID-related deaths are attributed to the age group above 90, 7.5% to 80-89, declining to 0.04% for age 40-49 and 0.01% for age 30-39, the latter more than an order of magnitude lower than previous estimates. We observe that the IFR traces the Yearly Mortality Rate (YMR) above ages of 60 years, which can be used as a model to estimate the IFR for other populations and thus other regions in the world. We predict an IFR lower bound of 0.5% for NYC and that 27% of the total COVID-19 fatalities in NYC should arise from the population below 65 years. This is in agreement with the official NYC data and three times higher than the percentage observed in Lombardia. Combining the PFR with the Princess Diamond cruise ship IFR for ages above 70 we estimate the infection rates (IR) for regions in Italy. These peak in Lombardia at 26% (13%-47%, 95% c.l.), and for provinces in Bergamo at 69% (35%-100%, 95% c.l.). These estimates suggest that the number of infected people greatly exceeds the number of positive tests, e.g., by a factor of 35 in Lombardia.^*^

## Introduction

The COVID-19 pandemic is one of the most important challenges facing the world today. Despite the large number of infected individuals and confirmed deaths, large uncertainties on the properties of the virus and the infection remain. In this article we focus on Italy, one of the hardest-hit countries at the time of writing, with more than 220,000 confirmed cases and more than 30,000 attributed deaths^a^.

Several numbers in Italy present statistical peculiarities such as the Case Fatality Rate^b^ (CFR), which exceeds 10% for Italy^1^, and has led to early estimates of high mortality^3^. CFR is heavily affected by issues unrelated to the underlying disease, such as the extent of testing. A more stable quantification is the Infection Fatality Rate^c^ (IFR), the knowledge of which is paramount to guide the public health response. The IFR, along with the Population Fatality Rate^d^ (PFR), allows us to estimate the Infection Rate^e^ (IR) which estimates how wide-spread the diseases is in the society and which informs government response.

Estimating IFR and IR is challenging, both due to the limited testing (hence poorly known number of infections) and the uncertainty in the number of fatalities attributed to COVID-19. Official data accounts for those that have been tested, mostly in the hospitals. However there may have been other deaths that were not tested and went unrecorded, suggesting an underestimate of the death rate by the official COVID-19 numbers. In addition, the official COVID-19 death statistics can be complicated to extract, as most of the infected patients that die in hospitals also suffer from other co-morbidities.

Given the uncertainties in the official COVID-19 fatality rate, it is important to explore other paths for obtaining it. In this article we propose a Data Science based counterfactual analysis, where we compare the weekly mortality rate for Italian regions in the first 4 months of 2020 with a model prediction obtained from historical mortality rates for the same time of the year. The model accounts for historical year to year variability due to the fluctuations caused by seasonal effects. We attribute the difference between the true 2020 data and the predicted counterfactual as excess deaths due to COVID-19.

## Data

We use the total Italian mortality data (due to all cause) from the Italian Institute of Statistics (Istat). The dataset contains the total number of daily deaths for 6,866 towns in Italy for the period of January 1st - March 31st (Class-2 data) for years 2015-2020, and for 4,433 towns upto April 15th (Class-1 data). We use the daily dataset with mortality in 21 age groups: 20 between age of 1-100 and one bin for ages above 100. To reduce the statistical noise we combine the daily data into week-long periods and 10 age groups.

We combine the data from the different towns in the same region for our analysis^f^. Class-2 data has over 86% of Italian population and over 95% completeness for regions in northern Italy. We use it to estimate excess mortality from February 22-March 28. We assume the missing population is random and scale up the estimated mortality from this dataset in proportion to the ratio of the sum-total population of the towns in our dataset with the total regional population, as per the 2010 census. Class-1 dataset is over 70% complete for northern Italy and we use it to estimate the mortality from March 28-April 11. The scaling for this dataset is determined by matching the estimated excess mortality with that of more complete Class-2 dataset for the period of February 22-March 28. To estimate the fatality rates, we will primarily rely on the most complete region of Lombardia (Class-1 97%, Class-2 78% complete) and province of Bergamo (Class-1 98%, Class-2 78% complete).

We compare our numbers with the reported COVID-19 mortality^g^. We assume that the age distribution of COVID-19 mortality in every region is the same as the national distribution, except for Bergamo which provides the age-distribution data.

## Methods

We estimate the true mortality count due to COVID-19 by comparing the current mortality to a prediction derived from the historical mortality in different regions of Italy. Specifically, we construct a counterfactual for every region, i.e. the expected mortality count under the scenario that the pandemic had not occurred. It is the best prediction given the historical probability distribution of the death rate time series data, combined with the trend in the data before the beginning of the pandemic. This approach is superior to the averaging of historical data in that it can account for the trends that may be correlated in time. We then compare this counterfactual with the reported total mortality numbers for 2020 to obtain an excess death rate.

We treat the past years, 2015-2019, as control units and the current year 2020 as a treated unit. There are *N =* 5 control units of 14-week time-periods from Jan 1st to April 11th (*T =* 14). Since Italy reported its first death due to COVID-19 on February 22nd, a conservative estimate is that the pandemic of COVID-19 started the week of February 16th with respect to mortality, corresponding to *T*_0_ *=* 6.

Let *Y*_0_ *=* [*X*_0_, *Z*_0_] and *Y*_1_ *=* [*X*_1_, *Z*_1_] represent the matrix for the mortality in control units and treated unit, respectively, in the absence of any pandemic, where *X* and *Z* represent the pre- and post-February 16 blocks of the matrix. Then the shapes of different matrices are - *Y*_0_: *N* × *T*, *Y*_1_: 1 × *T*, *X*_0_: *N* × *T*_0_, *Z*_0_: *N* × (*T* − *T*_0_) and correspondingly for *X*_1_ and *Z*_1_. Since the treated unit undergoes a pandemic, we observe 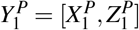 instead of *Y*_1_. Given the data from the previous years, *Y*_0_, and the current data, 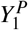, we are interested in predicting the counterfactual *Y*_1_ in the absence of pandemic. This can then be compared to the factual data 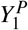 to assess the effect of pandemic.

In the simplest model, the expected the mortality count in 2020 is the mean of historical data 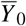. Thus

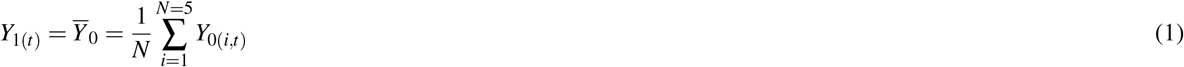

However, this is completely agnostic of the observed pre-pandemic data and ignores the time trends that may help improve the counterfactual. We improve on this model with two alternatives, a Conditional Mean with a Gaussian process (CGP), and a Synthetic Control Method (SCM).

CGP for the counterfactual analysis^4^ assumes a Gaussian distribution of the data and requires the knowledge of the kernel, which defines the covariance matrix of the data. Given the small size of control sample (N=5) as compared to the number of weeks (T=14), we adopt a kernel for the covariance matrix that combines a non-stationary kernel of the first few principal components (PCA) with a stationary component. We first estimate the principal components (*P*_1_*…P*_5_) of our control units *Y*_0_ for every region, finding that the first 2 explain more than 90% of the variance in the control data and hence do a 2 component PCA kernel. We add a stationary squared exponential kernel and determine its amplitude and length-scale from the data. This choice provides a good trade-off between capturing the variations in the data while avoiding over-fitting. We assume no noise, so by construction CGP predictions go through the pre-pandemic data points. The associated data covariance matrix is 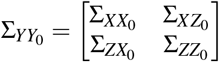.

The counterfactual *Y*_1_ follows the same distribution as the control units, i.e. a multivariate Gaussian with mean 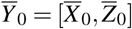 and covariance 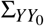. We are interested in the prediction of post-pandemic *Z*_1_: the conditional mean given the pre-pandemic data 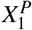 and the post-pandemic control mean 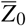 is

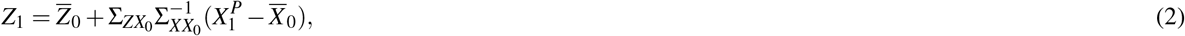

and the corresponding covariance matrix is

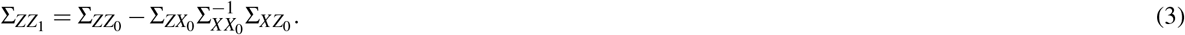

We use the diagonals of this covariance matrix as the error on the predicted counterfactual.

**SCM**^5^, our second approach, is a data driven method with minimal assumptions regarding the underlying data distribution. It estimates the counterfactual of the treated unit as a weighted combination of control units. The weights for various control units are estimated by minimizing the difference between the counterfactual and the observed data for the pre-pandemic period. Thus if W is the weight vector for the control unit, then we minimize

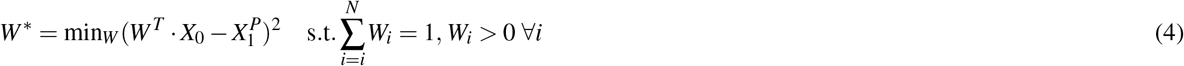

We have assumed a Gaussian, feature independent noise for the pre-pandemic data and put a positivity and unit L_1_ norm constraint on the weights. Given these weights, the counterfactual is predicted as

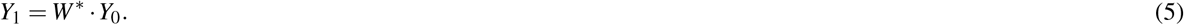

## Results

We begin by showing the counterfactual predictions for all the regions in the country in Figure 1. We plot predictions from both, SCM and CGP. For comparison, we also show the historical 2015-2019 data and their mean. We note that the SCM and CGP methods both trace the pre-pandemic data closely (the latter by construction). However, the historical mean estimates are generally higher. The mortality in Italy has been below-average in the first 2 months of 2020, probably due to a milder than usual flu season. This discrepancy demonstrates why SCM and CGP provide better counterfactuals than the historical mean: they take into account this years pre-pandemic mortality and exploit the time-correlations in the mortality rates to make more accurate and precise predictions. Despite this, for some regions like Lazio, the observed 2020 mortality is on the tails of the distribution represented by control units and hence our counterfactuals do not provide good fit. In these cases, and where our predicted excess mortality is lower than the reported COVID-19 fatalities, we will use the latter for our estimate of COVID-19 deaths. It only makes a statistically significant difference for the region of Lazio and age-groups below 30 years of age in few other regions, but otherwise does not affect our conclusions with any significance. Figure 1 shows a clear excess in mortality over the counterfactual predictions after the week ending on Feb 22, when the first COVID-19 related deaths were reported in Italy. This excess is primarily seen in the Northern regions which are the hardest hit. In the remainder of this work, we focus on these regions and the province of Bergamo (see also^2^ for an earlier analysis).

**Figure 1.**
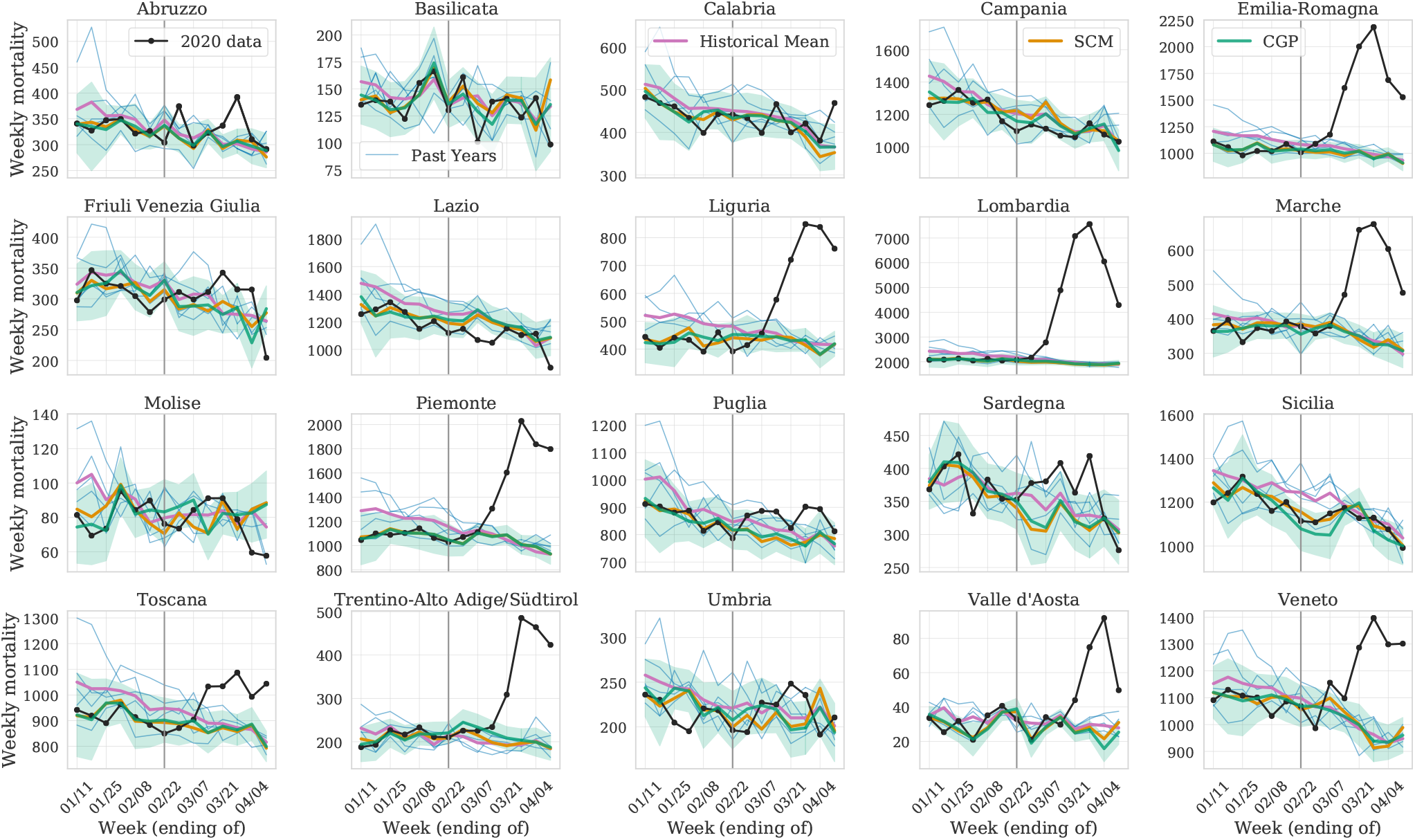
Validating counterfactuals for the pre-pandemic data. we show the observed weekly mortality due to all causes for the period of January 1 - April 11 (black) in all 20 regions in Italy, and our prediction for the expected mortality in the absence of COVID-19 (conditional Gaussian Process (CGP) with its 1*σ* confidence interval in green and synthetic controls method (SCM) in orange). The first reported COVID-19 mortality occurred in the week ending on February 22 (gray vertical line). The historical data from 2015-2019 (blue) and corresponding historical mean (pink) is shown for comparison and is not a good fit to the observed pre-pandemic data.

In Figure 2, we show the excess deaths over the expected counterfactual for every week of reported data. Figure 2a shows that the excess weekly mortality is significantly higher than the official COVID-19 deaths in all regions, right from the beginning of the pandemic. Figure 2b shows the cumulative excess in mortality compared to the total reported COVID-19 deaths at the end of each week. As of April 11, we find that the worst affected states such as Lombardia and Emilia-Romagna have likely under-estimated the mortality by factors of 1.5. while other regions like Piemonte and Toscana have under-estimated mortality by a factor of 2.

**Figure 2.**
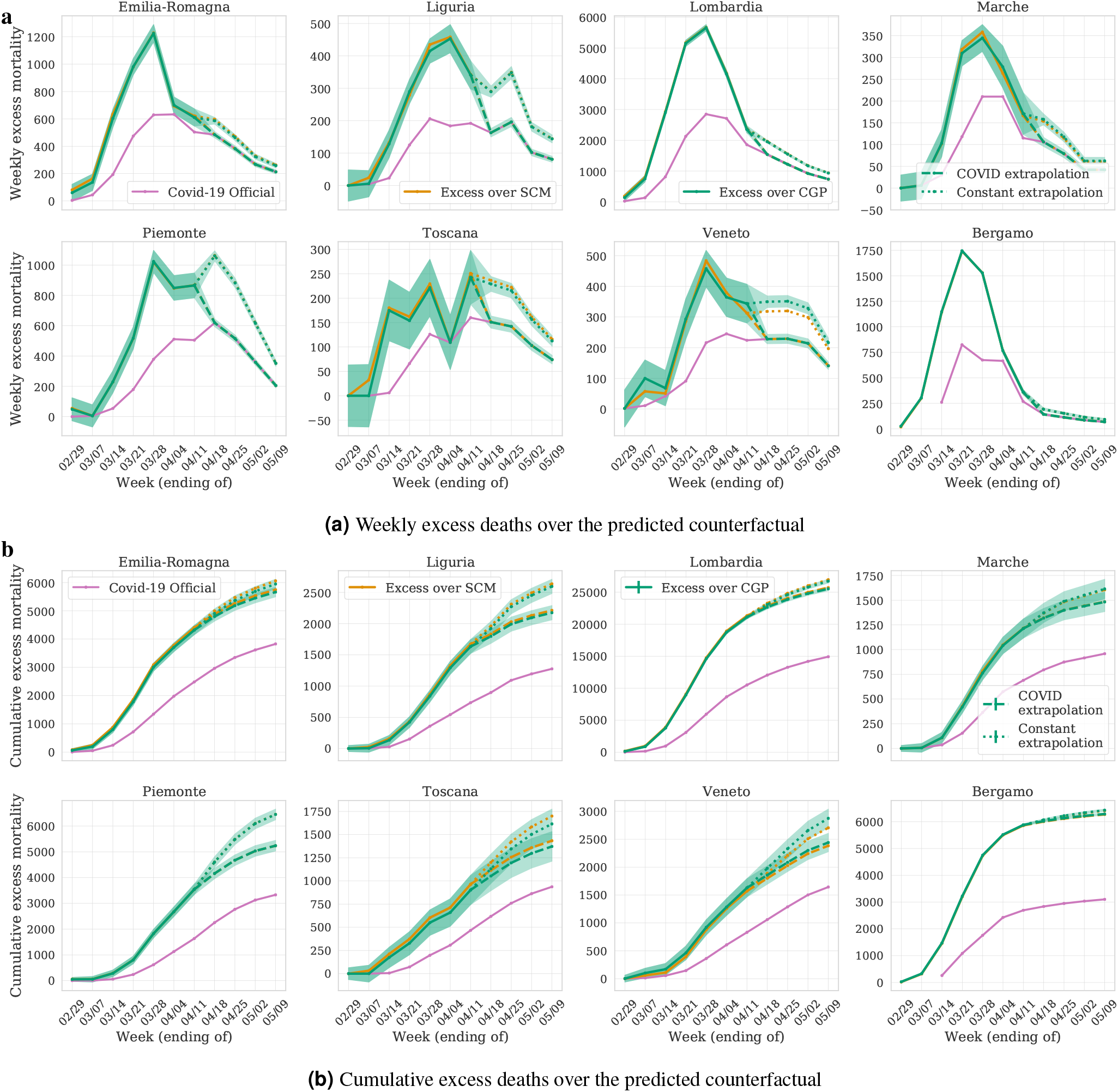
Excess mortality compared to reported COVID-19 deaths in regions of Northern Italy and the province of Bergamo. **a**, excess weekly deaths, and **b**, cumulative excess deaths, over the predicted counterfactual in comparison to the reported COVID-19 deaths (in pink) for the period since February 23rd (available COVID-19 data). Estimates from both the counterfactuals, SCM (orange) and CGP (green) agree. We find that COVID-19 deaths are under-reported by multiple factors for every period and every region. We extrapolate the data excess beyond April 11th with dashed-lines. To do this, we make the conservative assumption that the ratio of excess deaths to COVID-19 reported deaths is 1 after April 18th.

In Figures 2 and 3 we have extrapolated our estimated excess from April 11 to May 9th under two different assumptions - i) a conservative assumption that the weekly excess mortality is the same as the reported COVID-19 deaths on April 18 and beyond (dashed lines). ii) a constant ratio assumptions: where we assume that the ratio of excess deaths to officially reported COVID-19 fatalities remains the same as on April 11 (dotted lines). We observe that the ratio of weekly excess to reported COVID-19 deaths is decreasing with time in all the regions, as is expected due to increased testing (see Figure 4). As of April 11 (Figure 2), the lowest ratio is observed in Lombardia with a value of 1.27. We thus expect i) to be conservative extrapolation giving a lower-bound on the total deaths, while ii) to give an upper-bound. Based on the two extrapolations, we estimate that the number of COVID-19 deaths in Italy is between 49,000 – 53,000 as of May 9th 2020, more than a factor of 1.5x higher than the official number. In the remainder of the paper, we will only use the conservative extrapolation to estimate the age-dependent and population fatality rates and infection ratios.

**Figure 3.**
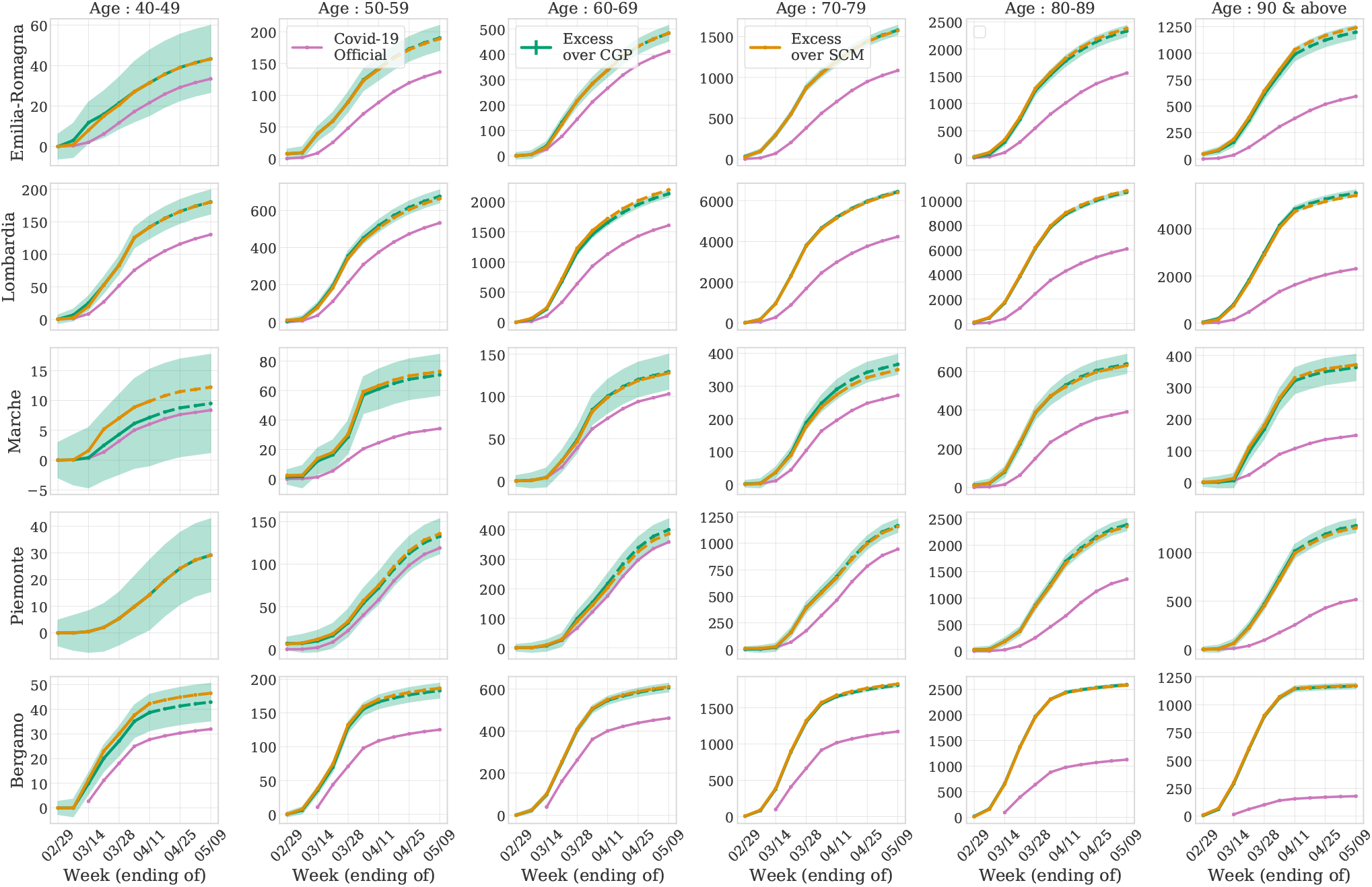
Age distribution of excess mortalities. Same as Figure 2b but for different age groups. We find a statistically significant excess over the reported COVID-19 deaths that is increasing with age.

**Figure 4.**
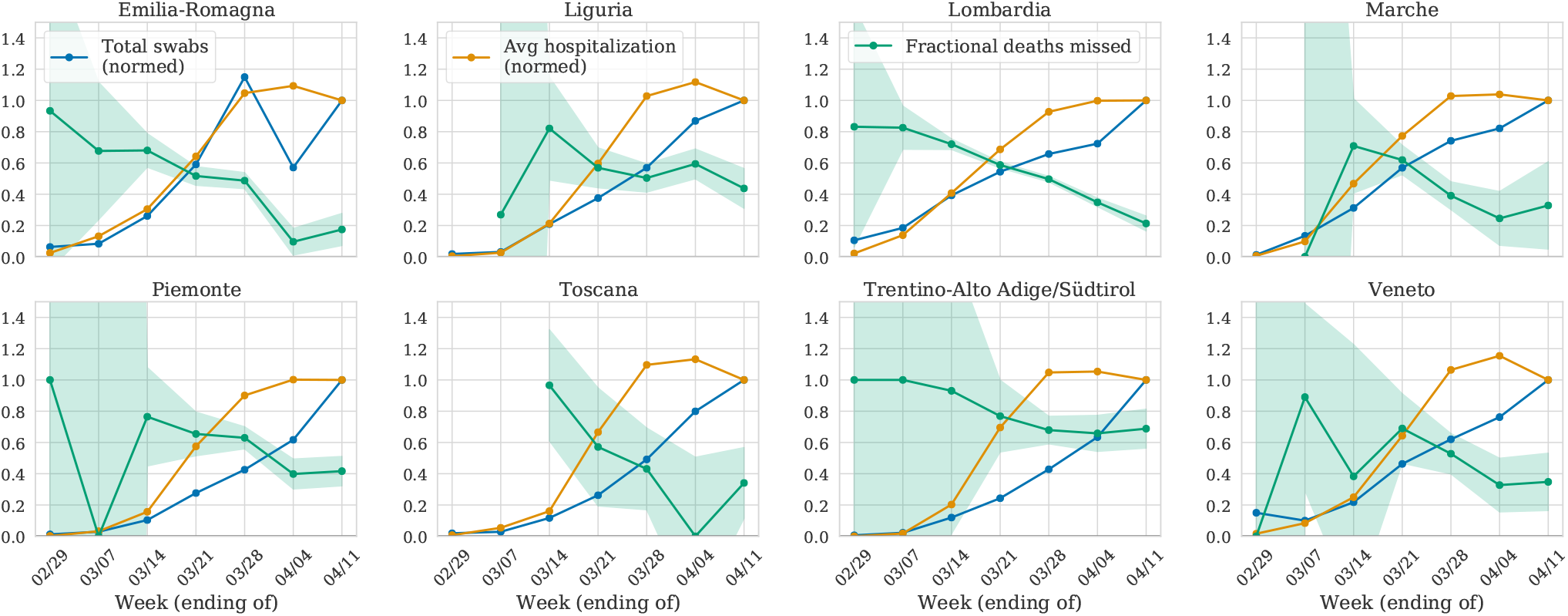
Fraction of missed deaths over time. For the period of the pandemic, we show per week the fraction of missed deaths (green), the number of hospitalizations (normalized as fraction of hospitalizations in week of April 11, in orange) and the number of tests conducted (normalized as fraction of tests conducted in week of April 11,in blue). We find that the missed fraction goes down as the number of tests increases while the hospitalizations have remained consistently high in the last 4 weeks.

In Figure 3 we show the excess mortality for different age groups in bins of 10 years above the age of 40. We find some agreement between the estimated excess and the reported COVID-19 deaths below the age of 70, but observe a significant and increasing discrepancy for higher age groups. This seems to suggest that testing and consequently probably also treating has been more complete for lower age groups.

### Attributing excess deaths to COVID-19

We attribute the excess deaths entirely to COVID-19. To back this assumption, we establish a correlation with COVID-19 with a regression analysis: We perform a 2-parameter fit between the official number of daily deaths attributed to COVID-19 and the daily excess deaths over the counterfactual, allowing the former to be scaled and shifted. We infer the time-lag and amplitude by minimizing *χ*^2^. We obtain best fit time-lags of −6 ± 0.5 days for Lombardia and −4 ± 1 days for Emilia-Romagna, and consistent, but noisier, results from other regions. The inferred time-lag suggests that the official COVID-19 mortality lags behind the total mortality, possibly a consequence of hospital treatment postponing death on average by several days.

However correlation is not causation and attributing the excess death rate to COVID-19 is still a strong assumption. Hence we discuss possible caveats. COVID-19 has put an enormous pressure on Italy’s medical system and social services. This could have led to fatalities that could otherwise be averted, causing us to overestimate the COVID-19 deaths. However, the pressure on the medical system is regional and likely sustainable for regions with a low number of official COVID-19 deaths, like Piemonte and Liguria. Instead, we consistently find a very large excess in mortality in most of the regions in Italy, including those that reported nearly zero COVID-19 deaths.

The temporal trend also lends a similar argument: the societal and medical systems should function normally in the earliest stages of the pandemic and get increasingly stressed as the number of infections increases. We see that the fraction of deaths missed by the reported COVID-19 fatalities is the highest in the early stage, and decreases as the number of reported infections increases. We show this in Figure 4 where we compare the fraction of deaths missed every week with the number of COVID-19 reported hospitalizations (normalized as the fraction of the hospitalizations in week of April 11).

Our hypothesis is that the excess deaths over official COVID-19 deaths are primarily due to the lack of testing in the initial stages of the pandemic. In Figure 4 we also show the number of tests conducted in every week as the fraction of tests conducted in the week of April 11. The trend supports our assertion that with an increase in testing as the pandemic evolves, the reported fatalities due to COVID-19 slowly catch up with the true current mortality and the increased pressure on medical systems did not have a statistically significant effect on the mortality.

There are also arguments that suggest we may have underestimated the COVID-19 death rate. Italy has been under lockdown since March 9, which may have reduced fatalities due to other common causes such as road and workplace accidents, or criminal activities. This can be studied by observing the death rate correlations with the lockdown data in regions with little or no infection, such as south Italy. There are several regions that do not show excess death rate, but none of them show a deficit death rate post March 9, so we assume that this effect is negligible.

### Fatality and Infection Ratios

Having established that the observed excess deaths can reasonably be attributed to COVID-19, we can use our estimates and uncertainties of the excess mortality from the CGP^h^ counterfactual to calculate the fatality rates and infection fractions for Italian regions. The left panel of Figure 5 shows the PFR in different age groups, the total number of excess mortality deaths attributable to COVID-19 as a fraction of the population. We find a steep age dependence of PFR: in Bergamo province, 1.81%, 4.67%, and 10.6% of the entire population in the age groups 70-79, 80-89, and 90+, respectively, died. For the entire population, the PFR is 0.58%. Since the PFR also corresponds to IFR if the infection fraction is 1 (maximum possible), we expect these to the be the most conservative lower limits on the (age dependent) IFR (Table 2).

**Figure 5.**
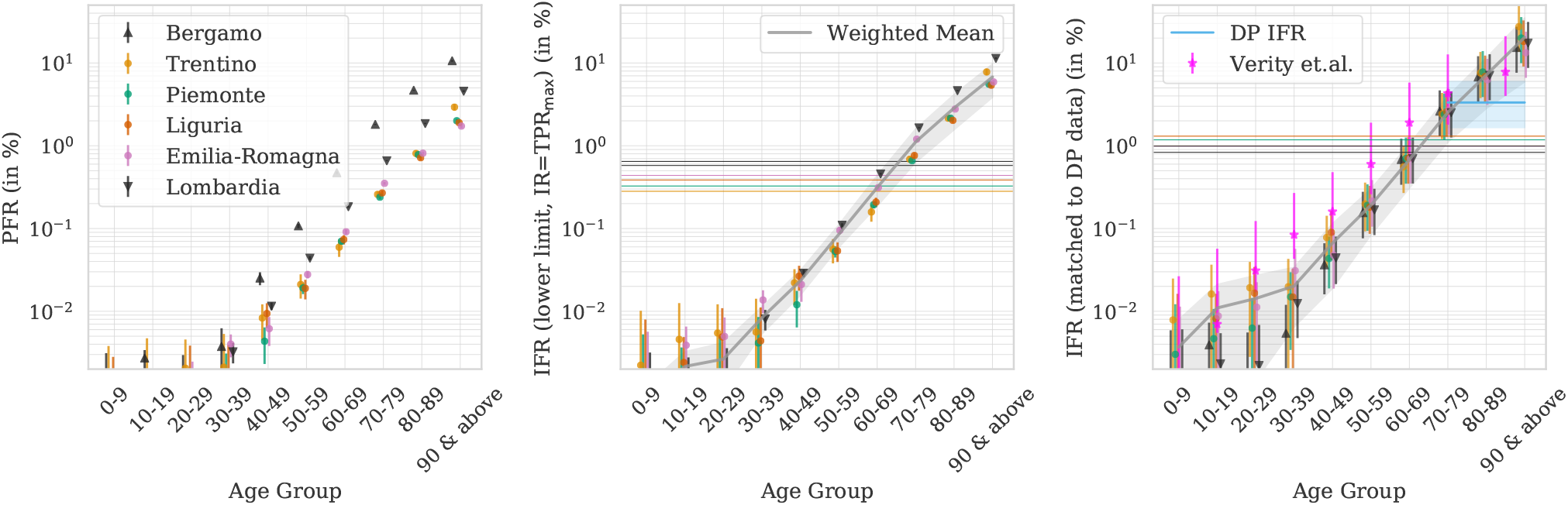
Fatality Rates for different age groups and regions. (Left) Population Fatality Rate (PFR) from the cumulative estimates divided by the regional population. (Center) Lower bounds on Infection Fatality Rate (IFR) using the maximum Test Positive Rate (TPR) as an upper bound on infection fraction. (Right) Estimates of the true IFR when normalizing the age 70-89 group to the Diamond Princess IFR (in shaded blue, with the corresponding Poisson error estimate). We also show estimates from Verity et al.^6^ in magenta, which give less steep age dependence. In center and right panel, the gray lines are weighted mean estimates for IFR with 1-sigma weighted standard deviation bands. The horizontal lines are the age-averaged IFR for the entire population. In all panels, we have staggered the points horizontally for every age group for better visibility.

#### Lower Limits on IFR

The central panel of Figure 5 shows the lower bounds on the IFR. To estimate the IFR from the PFR, we need the infection ratio (IR) of the population. Here we have used the Test Positivity Rate (TPR), the fraction of positive to total tests, as an estimate of the fraction of infected population. Due to lack of testing and the criterion of primarily testing people with symptoms, this should be an upper bound on the IR in the early stages of the pandemic. For every region, we use the maximum of cumulative TPR estimated up to May 2nd as our estimate of IR. This should be an upper-limit on IR and hence give a conservative lower bound on the IFR. We further assume that this ratio is age independent in every region^6^. The age averaged lower bounds on IFR are shown in Table 1, with the most robust estimate of 0.65 ± 0.02% IFR lower bound from Lombardia, consistent with 0.58% lower bound from Bergamo province.

**Table 1.** Estimated fatalities, Infection Rates (IR) and IFR. We estimate total deaths (as of April 11), lower limit IFR (by assuming IR=TPR for all regions except Bergamo, for which we take IFR lower bound=PFR) and IR by normalizing to DP IFR for age group above 70 years. Completeness is the fraction of regional population for which we have mortality data in our main dataset. The Total Death errors are 1 sigma errors (68% cl), and 95% cl for IR from DP.

| Region | Population<br>(in<br>millions) | COVID-19<br>(reported<br>deaths) | Completeness<br>(available data) | Total Deaths<br>(predicted) | TPR<br>(max) | IFR in %<br>(lower<br>limit) | IR from DP<br>mean (95%ci) |
| --- | --- | --- | --- | --- | --- | --- | --- |
| Emilia-Romagna | 4.41 | 3827 | 0.95 | 5662 ± 166 | 0.29 | 0.44 | 0.13 (0.06-0.23) |
| Liguria | 1.60 | 1276 | 0.92 | 2175 ± 113 | 0.35 | 0.39 | 0.10 (0.05-0.19) |
| Lombardia | 9.86 | 14924 | 0.97 | 25530 ± 286 | 0.40 | 0.65 | 0.26 (0.13-0.47) |
| Marche | 1.57 | 958 | 0.83 | 1481 ± 97 | 0.43 | 0.22 | 0.09 (0.04-0.16) |
| Piemonte | 4.43 | 3331 | 0.93 | 5237 ± 204 | 0.36 | 0.33 | 0.10 (0.05-0.18) |
| Toscana | 3.73 | 937 | 0.89 | 1372 ± 158 | 0.18 | 0.21 | 0.03 (0.02-0.06) |
| Trentino/Südtirol | 1.05 | 438 | 0.92 | 1107 ± 74 | 0.38 | 0.28 | 0.11 (0.05-0.19) |
| Veneto | 4.93 | 1643 | 0.87 | 2440 ± 164 | 0.09 | 0.54 | 0.05 (0.02-0.09) |
| Bergamo | 1.09 | 3101 | 0.98 | 6285 ± 39 | 1.00 | 0.58 | 0.69 (0.35-1.24) |

#### IR and IFR calibrated on Diamond Princess

The PFR can also be combined with an independent estimate of the IFR to obtain the Infection Rate (IR), IR=PFR/IFR. At the time of writing, the only large dataset with complete testing and hence unbiased estimate of the IFR is the Diamond Princess (DP) cruise ship. For our analysis we assume that the age dependent IFR is location independent: we account for age differences, but not for other differences between the DP and Italian populations in the same age group.

As of April 18, 11 out of 330 DP infections in the age group above 70 had been fatal (a few of the fatalities do not have age information). This results in an IFR for this age group of 3.3% and we assign Poisson error distribution to this. The population distribution in this age group on the DP was 80% in 70-79 and 20% above 80^7^. For each region of Italy, we re-scale to match this age distribution and hence match the age-weighted IFR to the DP in the 70-89 age group. Combining this with the corresponding PFR, we are able to estimate IRs for this age-group. Under the assumption of age-independent IR, we can then also derive IFR for all the other age-groups (Table 1). IR range from 3% up to 26% (13%-47% 95% cl) in Lombardia and 69% (35%-100% 95% cl) in the province of Bergamo. In all cases the estimated mean IR is below the upper limit set by the maximum TPR.

#### Age dependence of IFR

The right panel of Fig. 5 shows our estimate of these DP-anchored IFR estimates. Since we make the assumption of constant IR for all age-groups, we focus on the regions with high IR (more than 10%) where this assumptions is more likely to hold. The most reliable data come from Lombardia and Bergamo, since they are nearly complete, past the peak, and has a high number statistics with small errors. The age dependent IFR range from below 0.04% for ages below 50 years to 2.53%, 7.12%, and 17.5% for ages 70-79, 80-89, and above 90 years, respectively, (Table 2). This is broadly consistent with the estimates from the Hubei province in China, but suggests a steeper age dependence as shown in Fig. 5 for Verity et al. analysis^6^.^3,6–9^. While the overall amplitude of our IFR estimates is anchored to DP, the relative age dependence of IFR is not.

**Table 2.** Age distribution of fatalities and IFR. We show the age-distribution of reported COVID-19 and our estimation of excess mortality for Lombardia, Bergamo and Emilia-Romagna, and the corresponding IFR estimates - the lower limit and estimated IFR from normalizing 70-89 IFR to DP princess data, as explained in the text. The errors are small for fraction of Total Deaths and IFR lower limit, and we report 95% for IFR from DP. We also show age fraction and yearly mortality for 2017: the latter traces IFR above age of 60 within 20%. Age averaged yearly mortality rate is 0.98% for Lombardia, 0.91% for Bergamo, and 1.13% for Emilia-Romagna. We also show Crude Mortality Rate per year, which traces IFR above age of 60 to within 20%.

| Region | Age group | Pop-<br>ulation<br>Fraction | Yearly<br>Mortality<br>Rate in % | Fraction of<br>COVID-19<br>(reported<br>deaths) | Fraction of<br>Estimated<br>Total<br>Deaths | IFR in %<br>(lower<br>limit ) | IFR in %<br>from DP<br>mean (95%cl) |
| --- | --- | --- | --- | --- | --- | --- | --- |
| **Lombardia**<br>Population IFR<br>(in % from DP<br>mean (95%cl))<br>0.99 (0.50-1.78) | 30-39 | 0.120 | 0.00 | 0.00 | 0.001 | 0.01 | 0.01 (0.00-0.02) |
|  | 40-49 | 0.161 | 0.11 | 0.01 | 0.007 | 0.03 | 0.04 (0.02-0.08) |
|  | 50-59 | 0.159 | 0.28 | 0.04 | 0.026 | 0.11 | 0.17 (0.08-0.30) |
|  | 60-69 | 0.120 | 0.75 | 0.11 | 0.083 | 0.46 | 0.70 (0.35-1.26) |
|  | 70-79 | 0.101 | 2.10 | 0.28 | 0.250 | 1.65 | 2.53 (1.26-4.52) |
|  | 80-89 | 0.060 | 6.60 | 0.41 | 0.417 | 4.65 | 7.12 (3.55-12.74) |
|  | ≥ 90 | 0.012 | 18.80 | 0.15 | 0.215 | 11.42 | 17.50 (8.72-31.33) |
| **Bergamo**<br>Population IFR<br>(in % from DP<br>mean (95%cl))<br>0.85 (0.42-1.49) | 30-39 | 0.120 | 0.00 | 0.00 | 0.001 | 0.00 | 0.01 (0.00-0.01) |
|  | 40-49 | 0.161 | 0.11 | 0.01 | 0.007 | 0.03 | 0.04 (0.02-0.07) |
|  | 50-59 | 0.161 | 0.26 | 0.04 | 0.029 | 0.11 | 0.15 (0.08-0.28) |
|  | 60-69 | 0.121 | 0.76 | 0.15 | 0.095 | 0.47 | 0.68 (0.34-1.22) |
|  | 70-79 | 0.094 | 2.10 | 0.38 | 0.283 | 1.81 | 2.61 (1.30-4.66) |
|  | 80-89 | 0.052 | 6.60 | 0.36 | 0.404 | 4.67 | 6.75 (3.37-12.07) |
|  | ≥ 90 | 0.010 | 19.30 | 0.06 | 0.182 | 10.60 | 15.31 (7.63-27.40) |
| **Emilia-**<br>**Romagna**<br>Population IFR<br>(in % from DP<br>mean (95%cl))<br>0.96 (0.49-1.77) | 30-39 | 0.116 | 0.00 | 0.00 | 0.003 | 0.01 | 0.03 (0.01-0.06) |
|  | 40-49 | 0.161 | 0.11 | 0.01 | 0.007 | 0.02 | 0.05 (0.02-0.09) |
|  | 50-59 | 0.157 | 0.29 | 0.04 | 0.032 | 0.10 | 0.21 (0.11-0.38) |
|  | 60-69 | 0.121 | 0.75 | 0.11 | 0.083 | 0.31 | 0.70 (0.35-1.26) |
|  | 70-79 | 0.103 | 1.99 | 0.28 | 0.270 | 1.20 | 2.71 (1.35-4.85) |
|  | 80-89 | 0.066 | 6.60 | 0.41 | 0.398 | 2.78 | 6.27 (3.12-11.22) |
|  | ≥ 90 | 0.016 | 19.10 | 0.15 | 0.205 | 5.90 | 13.33 (6.62-23.86) |

#### Crude Mortality Rate per year as a tracer of Infection Fatality Rate

The infection fatality rate even when stratified by age can still vary between different populations. One reason for this is varying prevalence of co-morbidities. We explore a simple model for generalizing our findings to different populations: In Table 2 we list the crude mortality rate per year (YMR) i.e. the fraction of the population that on average dies within a year for each age group and region. We find that the YMR traces the IFR for ages above 60 within 20%. Thus a useful expression to obtain crude estimates of the IFR lower bound, mean and upper bound (the former from Italy TPR, the latter two using DP, at 95% cl) is 0.8, 1.0, 1.8 times the Yearly Mortality Rate (YMR) of the given population (Table 2). This simple model can then be used to make predictions for the IFR in other populations.

Using this model for New York City which has population YMR of 0.62%, we obtain the lower bound (mean) on population IFR of 0.50% (0.62%). This is similar to our estimate from combining age-dependent IFR from Italy data. It is also consistent with a lower bound of 0.55% IFR estimated by combining the NYC COVID-19 PFR (including probable deaths) of 0.25% with a TPR of 0.45% as of early May, independent of the Italian data. From our YMR model, if we take 0.62% IFR estimate together with 0.25% PFR, we find that 40% of NYC residents are already infected. However this number decreases if the actual IFR is higher than 0.62%. As of April 22, with 9900 confirmed deaths, this IFR of 0.62% predicted 19% IR which was in good agreement with then estimated IR of 21% from seropositivity tests.

Using age-dependent YMR, we can also estimate the proportion of deaths in different age-groups. The proportion of YMR predicted deaths in age groups 45-64, 65-74 and above 75 is 19%, 18% and 55% respectively^i^. This is in agreement with the current official NYC COVID-19 death fractions of 22%, 25% and 50%^j^. Interestingly, these numbers predict the higher death fraction for younger population that is seen in US as compared to Italian regions like Lombardia where only 8% of fatalities are in age group less than 65 years of age.

## Discussion

To our knowledge, our analysis makes use of the most recent available mortality dataset with the highest completeness and best statistics to estimate fatality rates for COVID-19. Our results suggest that a significant population of older people has died of COVID-19 without getting tested and entering into official statistics. This leads to an underestimation of total deaths in Italy by more than a factor of 1.5.

For policy decisions, one of the key parameters is the IFR and in this article we derived a strong lower bound from combining the PFR with the TPR. This bound is 0.65% from Lombardia and 0.55% from NYC. On other hand, our bound of 0.58% from Bergamo is independent of IR and TPR since we make the most conservative assumption of IR ~ 1 to derive this.

Our work has implications on the age distribution of the mortality, which is skewed even further to the elderly population than the offical COVID-19 statistics suggest (see Table 2). Due to the high number statistics we can estimate the IFR in lower age groups very precisely: for example, we obtain 0.04% IFR (0.02%-0.07% 95 % cl) in age group 40-49, a lot lower than previous estimates^6^. At the same time, using our YMR model, we are able to get a first estimate of the proportion of fatalities in different age groups while accounting for regional age-distribution and health status of the underlying population. We show that this predicts the fraction of total fatalities attributed to the younger population (<65 years) in New York City to be 27% as compared to only 8% in Lombardia, a factor of three difference which is also observed in the official COVID-19 reported fatalities after taking into account probable deaths. It suggests that COVID-19 kills the weakest segments of population as tracked by the YMR.

For this work to be globally relevant, we can test the hypothesis that the age dependent IFR is location independent by comparing our lower bounds and mean-estimates to IFR estimates from other regions. We find that our IFR estimates are lower than the CFR estimates of most countries^k^, which is as it should be since CFR is commonly taken to be an upper bound on the IFR^l^. For countries like Iceland with significantly different age-distribution as compared to Italy, it is important to take these age-distributions into account to explain the observed differences. For instance, using our YMR model which inherently takes regional age-distribution into account, we estimate the IFR lower bound for Iceland to be 0.5%, as compared to the current Iceland CFR of 0.56% (0.25%-1.02%, 95% c.l.).

Another estimate of IFR to validate our lower-bounds comes from serology tests which estimate the IR and can be combined with the PFR. At the moment these tests suffer from the specificity error (false positive rate), which if not corrected can overestimate the overall IR. The largest serology survey to date has been performed in the Czech Republic, where the IR measured on a sample of 26,000 people was found to be only 0.4%. This could be an upper limit due to the poorly known false positive rate of the test. With the current PFR of 2.5 × 10^−5^ (which could be an underestimate to the true PFR if some deaths have been missed) this translates into IFR lower bound of 0.64%, very similar to the other lower bound IFR estimates in this paper (Bergamo, Lombardia, NYC). An example of a serology study where specificity is a small correction is the study in Gangelt^11^, which finds an IR of 14%, and converts it to a 0.36%-0.41% mean IFR based on 7-8 deaths. The Poisson distribution with 8 deaths gives 0.18%-0.81% IFR at 95% c.l., which means that the number of deaths should dominate the error. This error has not been taken into account by the analysis. The study could further be biased low by a possible underestimation of the total death count.

When comparing IFR across different regions, it is also important to note that - i) IRs can vary a lot within a single country (Table 1), ii) our assumption of age-independent IR might not remain valid. The best example for the latter comes from Singapore where the CFR is 0.1%. If it is taken as an upper bound to the IFR would violate our lower bounds on the IFR. However this contradiction likely arises from an in-homogeneous IR. In fact, anecdotal reports suggest that most of the infections are among the younger immigrant worker population, and the IR among the older population could be lower than the TPR. Thus age-dependent IR could also be a limitation to our analysis. We note, however, that such age-dependence is more likely for low IRs and our analysis has focused on regions with presumably high IRs of northern Italy. Furthermore, our age dependent PFR from the province of Bergamo give a lower limit to the IFR (Figure 5) which is independent of the IR.

In addition to being consistent with low CFR regions, our analysis also sheds light on the puzzle of high CFR in regions of Italy, for example 20% in Lombardia. This high CFR can be explained by the high IR. In Lombardia, the total number of administered tests as of May 9, 2020 was 477000, which is ≈ 5% of the population. With these tests, 0.8% of the population was tested positive, A comparison to our estimated 23% infection rate suggests that the infection rate is 35 times higher than the number of test positives. If tested cases are the most severe cases that likely required hospitalization, their fatality rate will be significantly higher than that of the overall infected population.

Our analysis relies on a few assumptions that we have highlighted throughout the text. Primarily, we attribute all the excess deaths to COVID-19 fatalities. The most direct way to verify this is to perform COVID-19 tests on every fatality, which is currently not done in any location. Alternative explanations for excess fatalities could partly be ruled out by repeating our analysis in other regions of the world. This approach is becoming increasingly feasible as data is made available for some locations (NYC, France, Spain) and preliminary analyses suggest a similar underestimation of COVID-19 deaths in other parts of the world. Some statistics such as those published by NYC are now including probable deaths in the official counts.

## Data Availability

We get the data from Istat - https://www.istat.it/it/files//2020/03/Dataset-decessi-comunali-giornalieri-e-tracciato-record.zip>
We make public, both the raw and clean datasets used at https://github.com/bccp/covid-19-data

## Acknowledgements

We thank Ehud Altman, Alex Krolewski, Zarija Lukic, Jasjeet Sekhon and Jacob Steinhardt for helpful comments and the Italian Institute of Statistics (Istat), and in particular Antonella Ciccarese, for their prompt responses and for making the data available on a short timescale.

## Author contributions statement

U.S., S. F., C.M. and V.B. designed the research and interpreted results. C.M. and V.B. did the main data analysis in consultation with U.S. and S.F.. S.F., C.M., G.S. and U.S. gathered datasets that C.M. and G.S. cleaned, and G.S. validated. All authors wrote and reviewed the manuscript.

## Footnotes

* We make our data and the analysis code public at https://github.com/bccp/covid-19-data. This repository will continue to get updated as more data becomes available.

a From: https://coronavirus.jhu.edu/map.html

b Defined as the ratio between COVID-19 attributed deaths and positive tests.

c Defined as the ratio between the number of deaths and the total number of infections.

d Defined as the ratio between the number of deaths and the total number of population.

e Defined as the fraction of population infected.

f The processed data used in this analysis is available at https://github.com/bccp/covid-19-data/tree/master/data/Italy, while the raw data is available (in Italian) at https://www.istat.it/it/files//2020/03/Dataset-decessi-comunali-giornalieri-e-tracciato-record.

g From https://github.com/pcm-dpc/COVID-19

h We have verified that the SCM method performs consistently.

i Using 2017 data from https://www.health.ny.gov/statistics/vital_statistics/2017/table32c.htm

j Data from https://www.l.nyc.gov/site/doh/covid/covid-19-data.page

k From: https://coronavirus.jhu.edu/map.html

l If only symptomatic cases are being tested and assuming a 50% asymptomatic ratio as suggested by the DP data, IFR<0.5 CFR. For countries with high test rates (e.g.Iceland) IFR<CFR may be more applicable.

